# Modelling the risk ecosystem of depression using machine learning in a population of young adults

**DOI:** 10.1101/2023.08.15.23294062

**Authors:** H. Fraser, A.S.F. Kwong, M. Brooks, B.I. Davidson, R. McConville, R. M. Pearson

**Author notes:** Joint authorship (R. McConville, R. M. Pearson).

## Abstract

Understanding what is predictive of early adulthood depression could help inform resource targeting and direction of approaches aiming to alleviate the personal, cultural, and economic burden of depression and similar disorders. This work uses multivariate longitudinal data (n=3487) measured from conception to adulthood from a UK based birth cohort of young adults (Avon Longitudinal Study of Parents and Children (ALSPAC)) and a machine learning approach to a) investigate whether episodes of early adulthood depression can be predicted from various risk factors across early life and adolescence, and b) interpret which factors are most important for predicting episodes of early adulthood depression. Here, we build four models to predict participants having an episode of early adulthood depression and show that the highest performing model can predict if people experienced symptoms of depression with an F1-score of 0.66, using a range of biological, behavioural, and early life experience related risk factors.

## Introduction

Two thirds of people who suffer from a mental health disorder in their lifetime will experience first onset by their early twenties ^1^, with adolescence and young adulthood shown to be a key developmental period associated with depression risk and vulnerability ^2–4^. Depression during this period is associated with poorer mental health outcomes in later life ^5^, higher health care utilisation and greater work impairment ^6^, and other adverse downstream social and health outcomes ^7^ that implicate this early-life transitional period as particularly consequential for future mental health. Understanding the determinants of mental health outcomes continues to be a core priority in mental health research, aligning with the understanding that depression is the result of a complex interplay of different factors.

Identifying mental health risk, and exploring methodological approaches to modelling this risk, may provide the framework for a multifaceted approach to understanding contributing factors to mental health disorders and their impact on society. Modelling risk provides a conceptual framework for prevention of symptom exacerbation and illness relapse and may help shed light on depression’s complex aetiology of genetic, psychosocial, and environmental influences ^8^.

There is a vast body of literature describing the varied array of risk factors for depression and mental health, implicating many areas of health, behaviour, lifestyle, and relationships as risk and prevention factors for future psychiatric outcomes. The majority of this work is based on separate studies and predominantly uses one statistical approach (regression modelling) which is limited in assessing accumulation or relative ranking of risk factors.

First we describe the current evidence base and justification for the risk factors included in this analysis. Depression is consistently reported as having a higher prevalence in women than in men ^9,10^, with meta-analyses showing that women are at significantly greater risk of both diagnoses and symptoms ^11^. Depression is suggested to have a significant genetic component ^12^ despite the architecture of genetic factors being complex. Multiple studies successfully identify genomic risk loci for depression ^13–15^, indexed by polygenic risk scores (PRS) for depression which can capture an individual’s risk for developing depression in the future ^16^, as well as being able to predict clinically relevant depression characteristics such as age of onset and severity ^17^. Family history of depression is proposed to have an associated intergenerational risk, transmitted through both genetic and environmental pathways ^18^. One recent study found that history of mental health problems was a strong predictor of depression in adolescent girls ^19^, with earlier research showing that having a parent with severe mental illness doubles the risk of developing a disorder of equal severity by adulthood ^20^.

Childhood trauma, such as experiencing emotional, physical or sexual abuse and neglect ^21^, is a well-known and evidenced precursor of adverse future depression in adolescence and adulthood, despite exact mechanisms being unknown. Studies of mental health outcomes after exposure to maltreatment during early childhood showed higher depression and post-traumatic stress disorder (PTSD) symptoms compared to those exposed to trauma at later developmental stages ^22^. One study found that within a cohort of chronically depressed patients, 75.6% had significant histories of childhood trauma ^23^. Childhood trauma is arguably not uncommon, with one study of adolescents in the United States indicating that 60% had experienced a ‘lifetime potentially traumatic event’ (PTE) ^24^. A recent UK based cohort study found that 31.3% of participants had reported childhood trauma, with 7.8% going on to experience PTSD by age 18 ^25^.

Models which incorporate longitudinal, life-course data benefit from insights into both the timing, severity and chronicity of early life traumas, and may be poised to answer questions on which aspects of health, lifestyles, relationships and behaviour in early life puts an individual at risk of depression in adulthood. It has been suggested that there is a ‘dose-response’ relationship between adverse childhood experiences (ACE) and psychiatric outcomes ^26^, with a higher dosage of trauma related to worse outcomes. There is also evidence to suggest that the type, timing and frequency of trauma may have differential effects on coping strategies and associated mental health problems ^27^. Reflecting on childhood development and caregiver dynamics, there is evidence to suggest that relationship health more broadly between parents / caregivers and children has a tangible impact on future depression, with feelings of being unloved or unwanted increasing the risk of depression in both men and women ^28^. Relationships with peers in adolescence and bullying behaviours has also been associated with adolescent depression and suicidality ^29^, with bullying and victimisation both preceding depression in adolescents as well as being a potential consequence of depression due to impacts on social functioning and self-esteem^30^. Research into social and romantic networks has also found a link between romantic relationship problems and future depression in a cohort of young people ^31^, indicating that support network strength may offset future depression risk.

Other relevant factors when considering depression risk include comorbidity with other illnesses ^32^, risk of depression in individuals with disabilities ^33,34^, and sleep management as both a symptom and modifiable target for treatment in depression ^35^. Socioeconomic environments such as financial hardship and indebtedness have been extensively researched ^36–40^, with increased financial resources suggested as a potential ‘active ingredient’ of socioeconomic approaches to combating youth anxiety and depression ^41^.

Alcohol use in adolescence has consistently been found to be associated with later depression outcomes ^42,43^, but worth noting that this relationship may also be comorbid and bidirectional ^3^. Adolescents with depression are also reported to have higher rates of substance use ^44^, which in turn is related to higher instances of suicidal behaviour ^45^. The relationship between cannabis use specifically and future mental health outcomes is not consistently agreed upon in the literature; whilst some older evidence suggests that depression and cannabis use do co-occur ^46,47^, other findings show that cannabis users were not more likely than non-users to be diagnosed with major depressive disorder (MDD) over their life time ^48^.

Given this extensive list of common and inter-related risk factors, it is an ongoing public health policy challenge to know which factors to prioritise when exploring and measuring risk. Such factors could be useful for predicting later depression and could inform more person-centred and targeted approaches to treatment and intervention, i.e., through integration into schools, community groups, family education and services. Understanding specific risk factors and how and when they map onto early-adulthood depression may also help guide decision making around types of intervention which may be most useful for whom. For example, focusing on improving and/or adapting lifestyle, or strategies to manage and deal with the impact of trauma.

This paper adopts a machine learning (ML) approach to modelling depression risk factors throughout child and adulthood, via longitudinal health data obtained from a large prospective longitudinal birth cohort of young adults based in the South-West of England. We train and evaluate a Random Forest (RF) ML model to predict if an individual will have at least one episode of depression by their mid to late twenties. We evaluate the predictive ability of the model using all available risk factor data, and then adopt a stratification approach which tests the predictive performance of models only trained on risk factors from specific domains (i.e., Biological risk, lifestyle factors, and childhood trauma and adverse experiences). This modelling strategy allows us to examine and measure risk factors for adulthood depression by incorporating a wide and extensive range of health and psychological data into a single model, which has very few limitations in terms of how much data it can utilise. This approach will allow us to examine a more comprehensive ‘ecosystem’ of depression risk, with the added benefit of not needing to state or make *a priori* assumptions of associations between variables, or manage multicollinearity between different factors ^49^ as required when adopting more traditional statistical approaches such as logistic regression.

The aims of this paper are threefold. We firstly aim to explore the utility of a RF ML model in a depression prediction task, using a wide range of different risk factors for depression. Secondly, we aim to understand the differential importance of each risk factor on predicting future depression mental health outcomes. Lastly, we discuss the interpretability considerations of our first two aims, with the overarching goal to examine the utility of these types of ML models as a tool in mental health research.

Table 1 shows the list of risk factors selected for classification, and their stratification into corresponding domains (‘All factors’ (AF); ‘Biological factors’ (BF); ‘Health behaviours and lifestyle’ (HBL); ‘Childhood trauma and adverse experiences’ (CTAE)). A schematic of these variables and dates of data collection in the ALSPAC resources can be found in Figure 1, indicating the temporal nature of the data and the time frame over which the risk factors have been measured and collected. A more detailed table describing ALSPAC data collection protocols and how variables used for analyses were derived can be found in the supplementary materials.

**Table 1:**
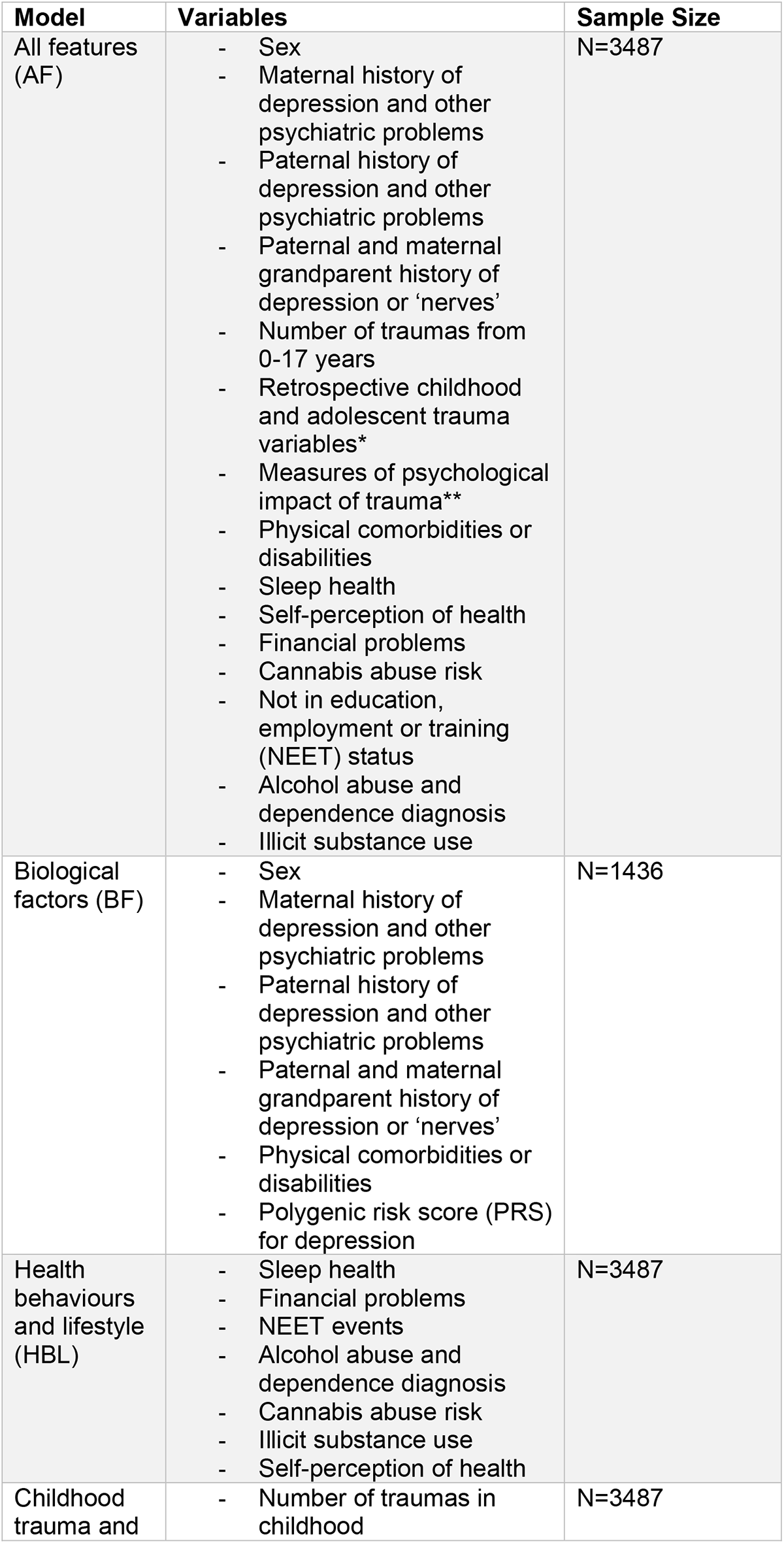
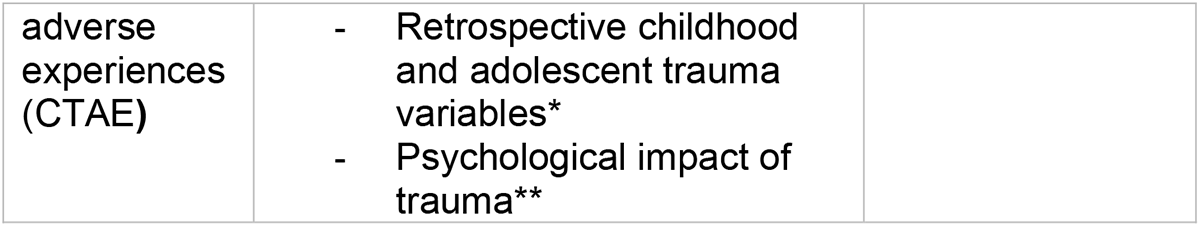
Description of models and variables used. (*Retrospective childhood trauma variables = feeling loved, domestic violence, sexual abuse, life-threatening accidents, sudden bereavement, witnessing death, controlling relationships, other extreme trauma or stress; ** Psychological impact of trauma variables = upsetting dreams, flashbacks, feeling bothered by reminders of experience, feeling jumpy / easily startled, paranoia).

**Figure 1:**
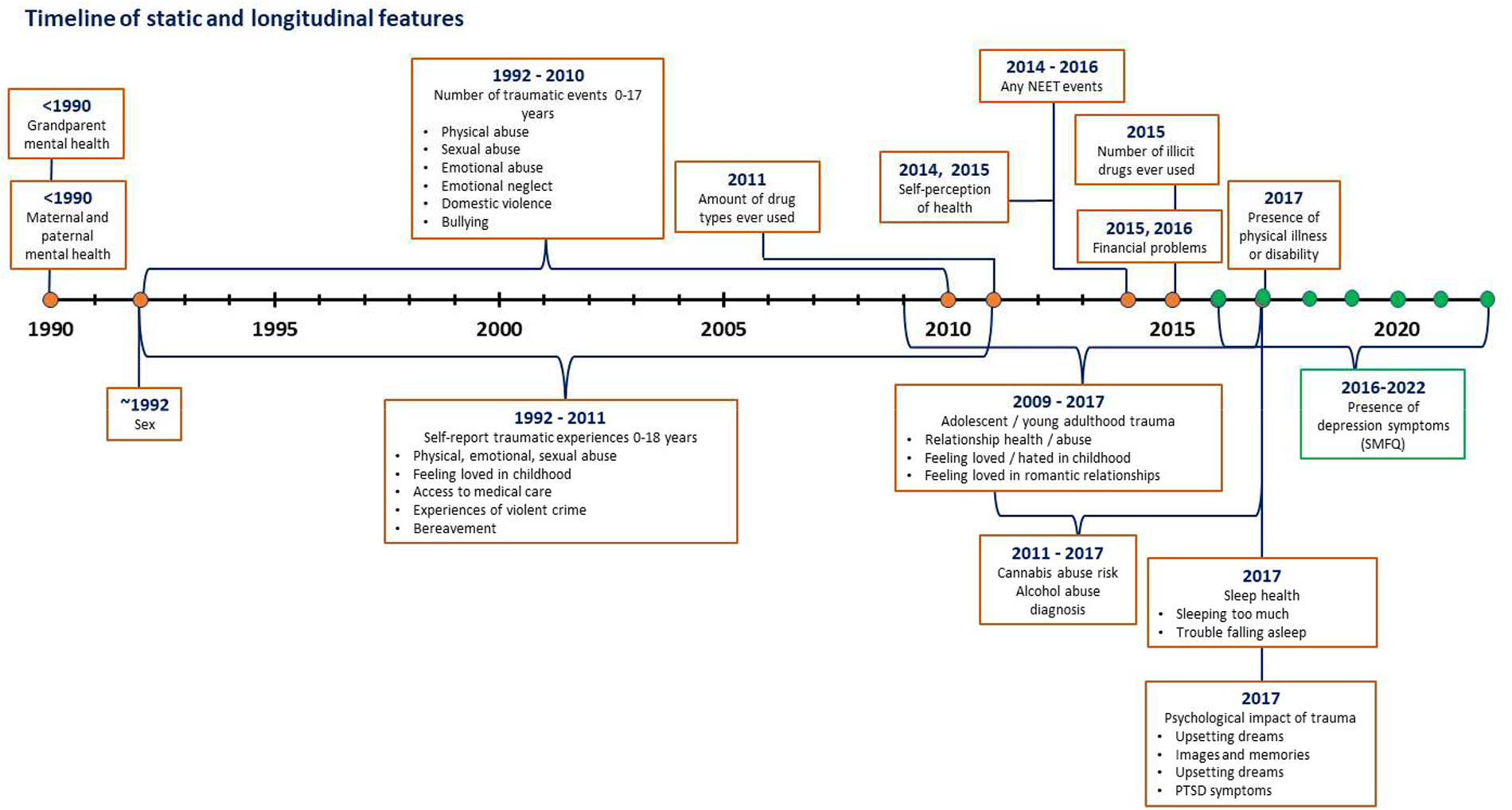
Schematic showing the timeline of longitudinal exposures modelling risk ecosystem in the ALSPAC resource [Post-Traumatic Stress Disorder (PTSD); Not in education, employment or training (NEET)

## Results

### Participant demographics

Table 2 in supplementary materials shows the demographic information for models AF, HBL and CTAE after data cleaning and exclusions (see methods). Table 3 in supplementary materials shows demographic information for Model-BF after data cleaning and exclusions. A separate sample of participants (n=1436) was used for Model-BF due to imputation limitations with the genetic data used in the biological features model. The sample for this model was therefore limited to participants who had complete case data.

### Machine learning results

Here, we present the results of four separate RF models (see Table 2) classifying early adulthood depression based on a variety of risk factors. Depression is defined in the context of this analysis as participants that have experienced at least one episode of depression between the ages of 23 and 28. An episode is defined as having a score of >=11 on the Short Mood and Feelings Questionnaire (SMFQ), a commonly used depression screening tool ^2,50^.

Each model was evaluated using four standard metrics to measure the models’ ability to classify both people with and without the experience of at least one episode of depression between the ages of 23 and 28. A table describing the metrics used to evaluate model performance can be found in the supplementary materials. The ‘depressed’ class was the positive class (target label ‘1’ if participant had experienced an episode of depression) within the analysis.

Model-AF had the best overall performance with a macro F1-score of 0.66. Whilst precision was relatively high for classifications of both depressed and non-depressed participants, a recall score of 0.44 in people with depression indicates that the model misclassified a large proportion of people in the model who had depression as not depressed. A higher F1-score for the non-depressed group (0.79) than the depressed group (0.52) indicates that the model performs better at classifying people without symptoms of depression in adulthood than people with symptoms.

Model-BF was the lowest performing model with a macro average F1-score of 0.55. Similarly to Model-AF, this model struggled to accurately classify people with depression (F1-score = 0.36) based on information from biological risk factors only.

Model-HBL was able to classify people with depression with a macro average F1-score of 0.60. This model outperformed Model-BF, but still suffered a performance decrement when classifying people with depression (F1-score = 0.45).

Model-CTAE was one of the highest performing models, with an F1-score of 0.65. Model CTAE was also the highest performing model when classifying people with depression, with an F1-score of 0.51.

To summarise, after training four separate models on depression risk factors, the model trained on all risk factors was the best performing model overall when predicting if someone will have an episode of depression in their early to mid-twenties. This was closely followed by the model trained on trauma risk factors. Biological risk factors in isolation did not perform highly when classifying depression.

**Table 2:**
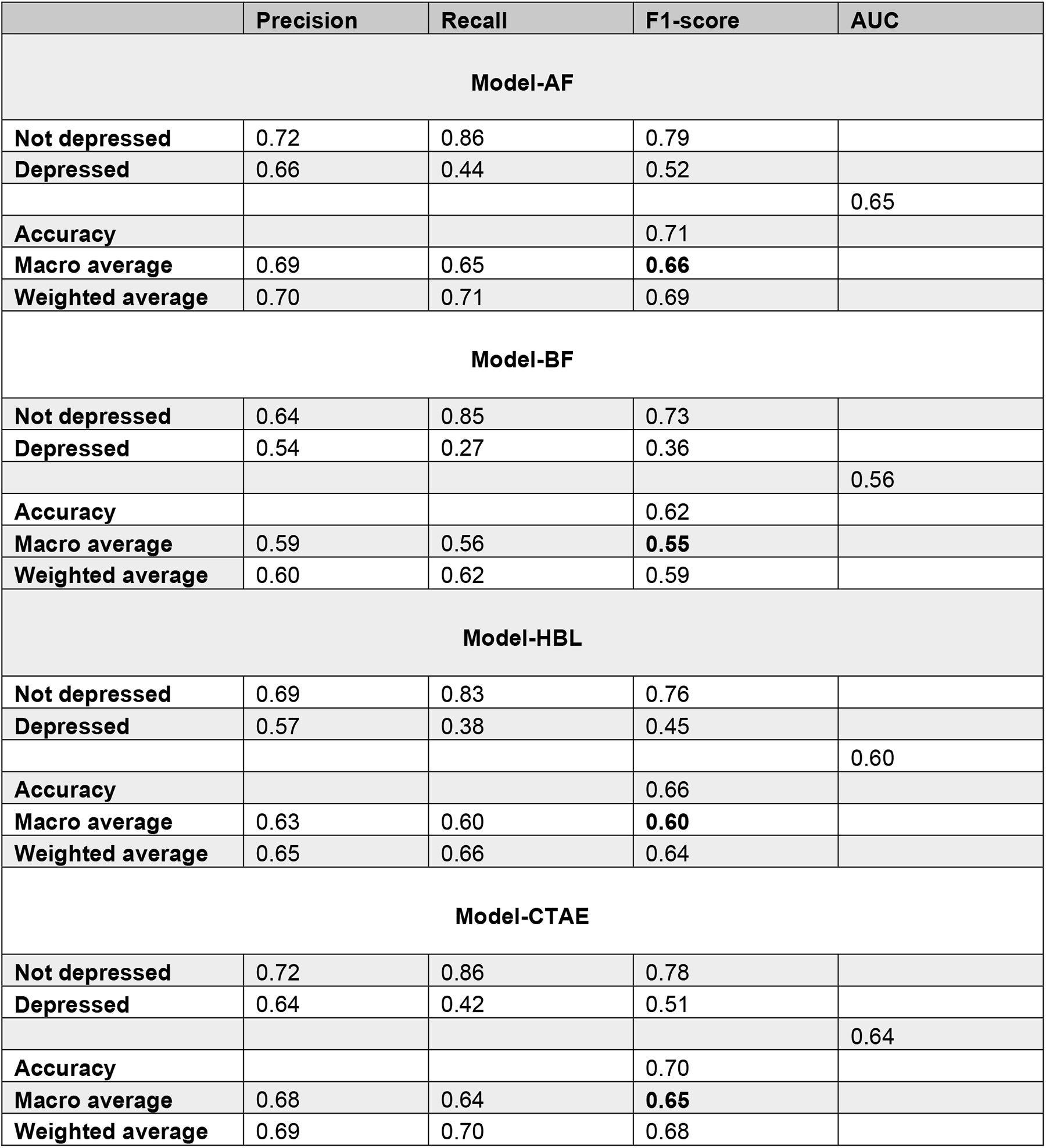
Performance evaluation of Models AF, BF, HBL and CTAE.

|  | Precision | Recall | F1-score | AUC |
| --- | --- | --- | --- | --- |
| **Model-AF** |  |  |  |  |
| **Not depressed** | 0.72 | 0.86 | 0.79 |  |
| **Depressed** | 0.66 | 0.44 | 0.52 |  |
|  |  |  |  | 0.65 |
| **Accuracy** |  |  | 0.71 |  |
| **Macro average** | 0.69 | 0.65 | **0.66** |  |
| **Weighted average** | 0.70 | 0.71 | 0.69 |  |
| **Model-BF** |  |  |  |  |
| **Not depressed** | 0.64 | 0.85 | 0.73 |  |
| **Depressed** | 0.54 | 0.27 | 0.36 |  |
|  |  |  |  | 0.56 |
| **Accuracy** |  |  | 0.62 |  |
| **Macro average** | 0.59 | 0.56 | **0.55** |  |
| **Weighted average** | 0.60 | 0.62 | 0.59 |  |
| **Model-HBL** |  |  |  |  |
| **Not depressed** | 0.69 | 0.83 | 0.76 |  |
| **Depressed** | 0.57 | 0.38 | 0.45 |  |
|  |  |  |  | 0.60 |
| **Accuracy** |  |  | 0.66 |  |
| **Macro average** | 0.63 | 0.60 | **0.60** |  |
| **Weighted average** | 0.65 | 0.66 | 0.64 |  |
| **Model-CTAE** |  |  |  |  |
| **Not depressed** | 0.72 | 0.86 | 0.78 |  |
| **Depressed** | 0.64 | 0.42 | 0.51 |  |
|  |  |  |  | 0.64 |
| **Accuracy** |  |  | 0.70 |  |
| **Macro average** | 0.68 | 0.64 | **0.65** |  |
| **Weighted average** | 0.69 | 0.70 | 0.68 |  |

### Comparison of model performances to baseline models

In order to validate the results of each model’s performance, we compared the performance of each model to a series of baseline classifier models which make classification predictions not based on the models’ input features. This comparison allows for an evaluation of whether each model is performing better than a model which has not been trained and provides a baseline performance level for each prediction model. Four baseline classifiers were used and are described in Table 3 below. For more information on each baseline classifier, see the following documentation for implementation in Python ^51^. Figure 2 shows each respective model’s performance when compared to each baseline classification model.

**Table 3:** Descriptions of the baseline classifiers used for model performance comparison.

| Baseline model | Prediction strategy |
| --- | --- |
| **Most Frequent** | Prediction method returns the class label of the highest frequency in the $y$ argument. |
| **Prior** | Prediction method returns the class label of the highest frequency in the $y$ argument (as above). |
| **Stratified** | The probability estimate randomly samples one-hot vectors from a multinomial distribution parametrized by the empirical class prior probabilities |
| **Uniform** | Generates predictions at random from the list of available classes observed in $y$, with each class having equal probability. |

**Figure 2:**
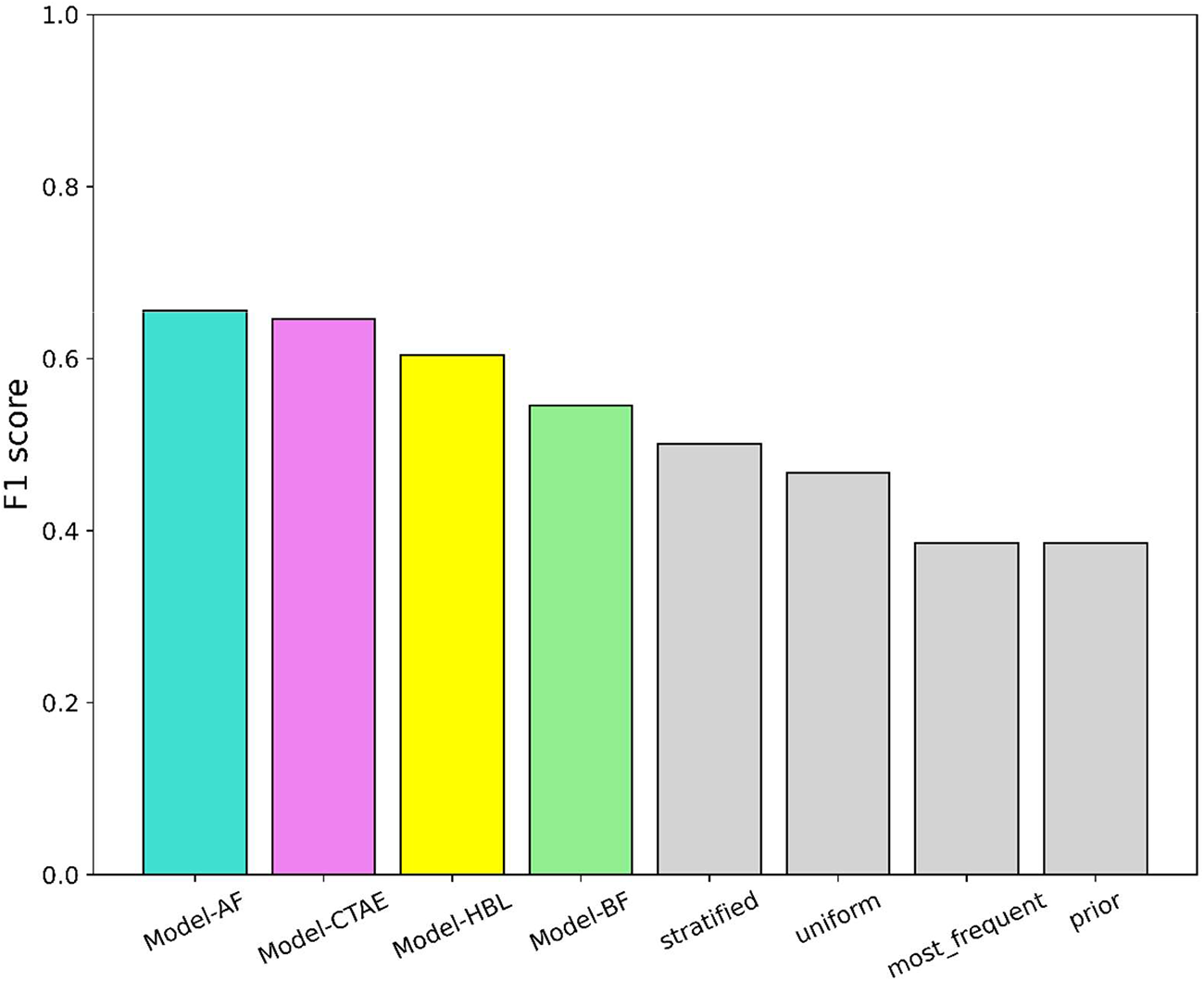
Performance of each model compared to untrained baseline classifiers.

### Permutation feature importance

Permutation feature importance is a strategy used to understand how much each of the models’ input features help the model to predict or classify by measuring the increase in prediction error after the input features have their values “permuted” (ie., shuffled or changed in value) ^52^. The higher the prediction error after a feature’s values have been permuted, the more important the feature is for the model’s prediction i.e. the model relies on the feature to make a prediction if permuting it increases error. Understanding the individual importance ranking for each feature helps interpretation of models in this context by facilitating the comparison of each risk factor’s importance. This allows for a hierarchy of risk to be quantified, which can be used to understand which risk factors should be explored in more depth.

Figure 3 shows that self-assessment of health, sleep problems in the past month at age 24, and feeling loved in a relationship are amongst some of strongest predictors of adult depression in Model-AF. There are high densities of variables from the childhood trauma and adverse events variable cluster in the top ten most predictive features, with variables relating to the psychological impact of traumatic experiences (feeling jumpy, experiencing flashbacks) suggesting a comorbidity between depression and PTSD / anxiety phenotypes which is supported in the literature ^53–55^. A key point to consider here is that whilst this ranking indicates the order of most predictive to least predictive variables, in this model none of the variables were highly strong independent predictors, with the highest importance ranking equalling 0.02.

Within the childhood trauma model, feeling loved when growing up is an important predictor of presence of depression episodes in adulthood, as well as the psychological burden of traumatic experiences (figure 6). Trauma in later childhood rated more important than early childhood trauma when predicting depression. The psychological burden of trauma (e.g., measures of feeling jumpy / easily startled) was more important to the model than specific traumas experienced. The scale of differences of feature importance in this model was also small, with the highest feature importance equalling 0.06.

**Figure 3:**
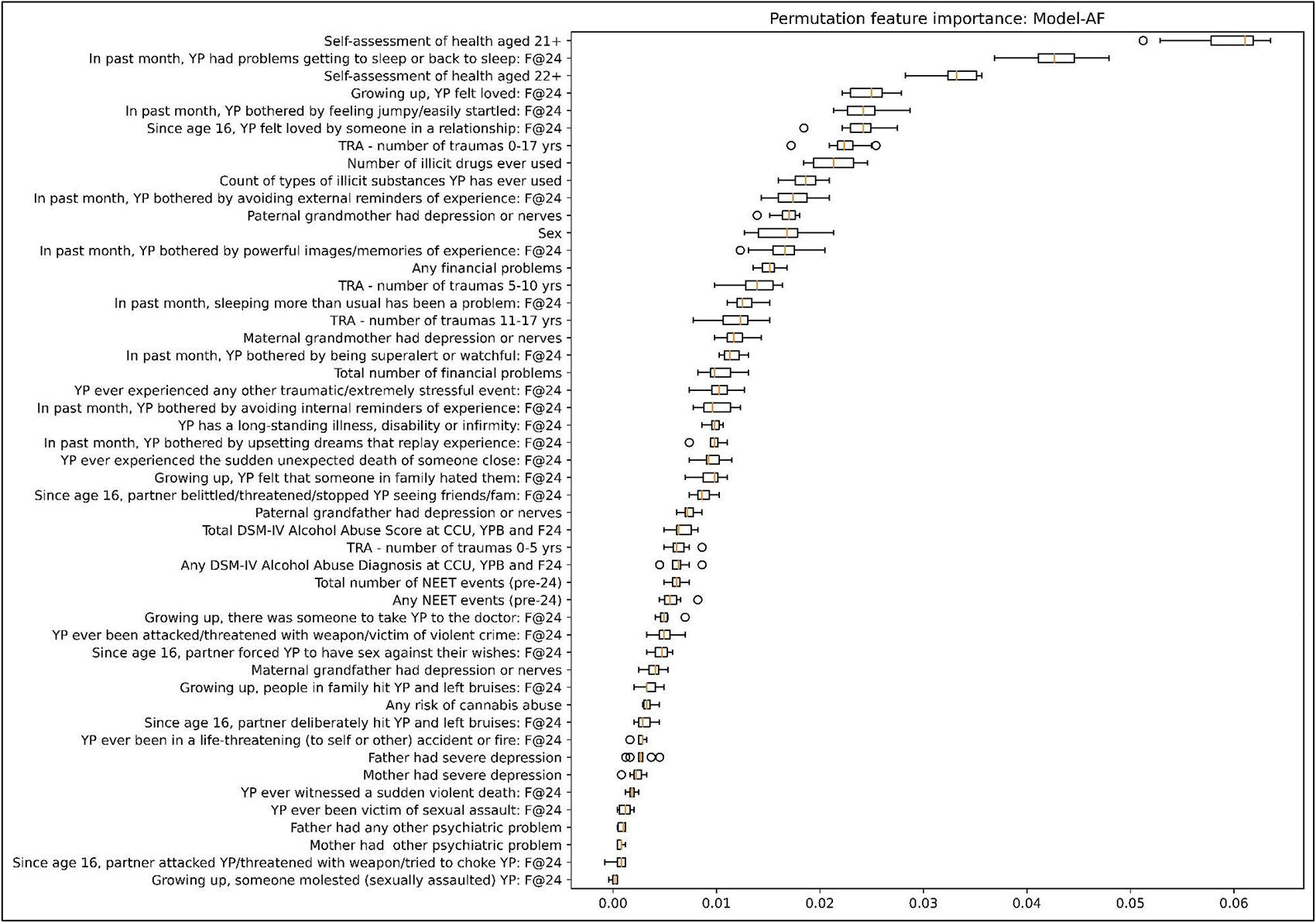
Permutation feature importance for Model-AF. Features ranked from most important (highest) to least important (lowest)

The polygenic risk score (PRS) for depression was the most predictive variable in Model-BF (figure 4), but care should be taken when interpreting this finding considering the low performance of Model-BF compared to the other trained models.

**Figure 4:**
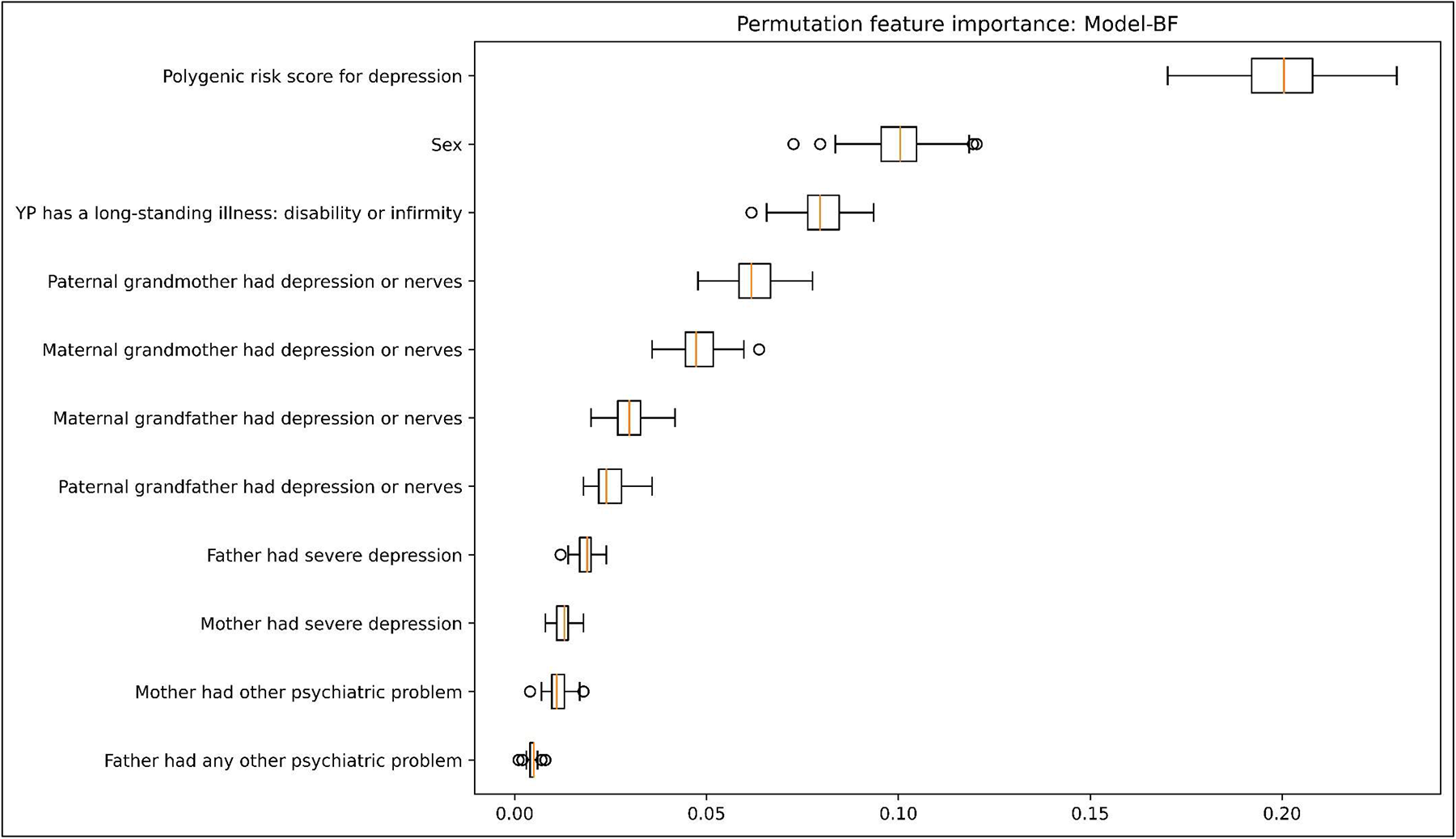
Permutation feature importance for Model-BF. Features ranked from most important (highest) to least important (lowest)

Self-perception of health and wellness and sleep health are amongst the strongest predictors of adulthood depression in Model-HBL (figure 5), ranking higher than other lifestyle features such as NEET events, financial problems, alcohol abuse and cannabis abuse risk.

**Figure 5:**
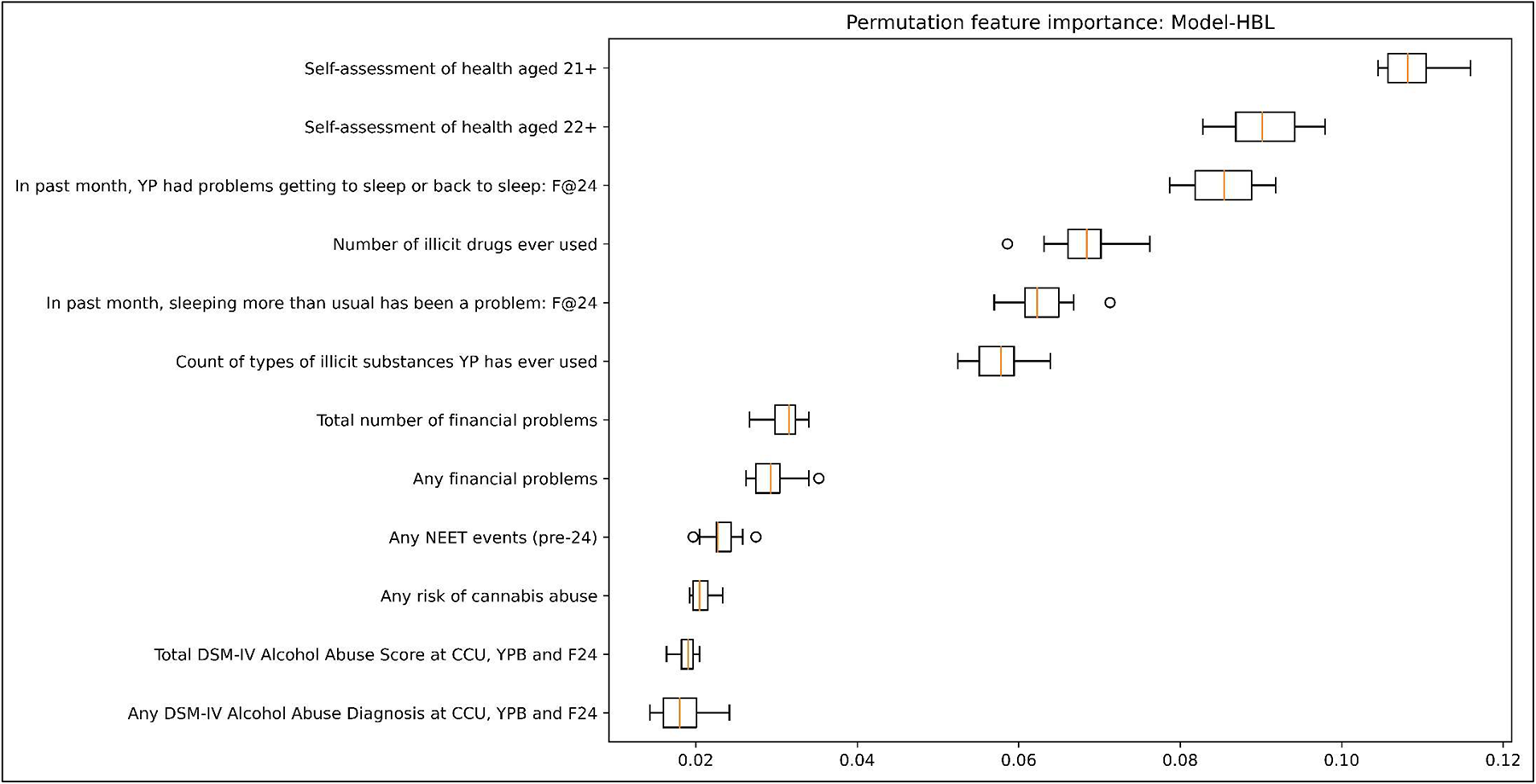
Permutation feature importance for Model-HBL. Features ranked from most important (highest) to least important (lowest)

**Figure 6:**
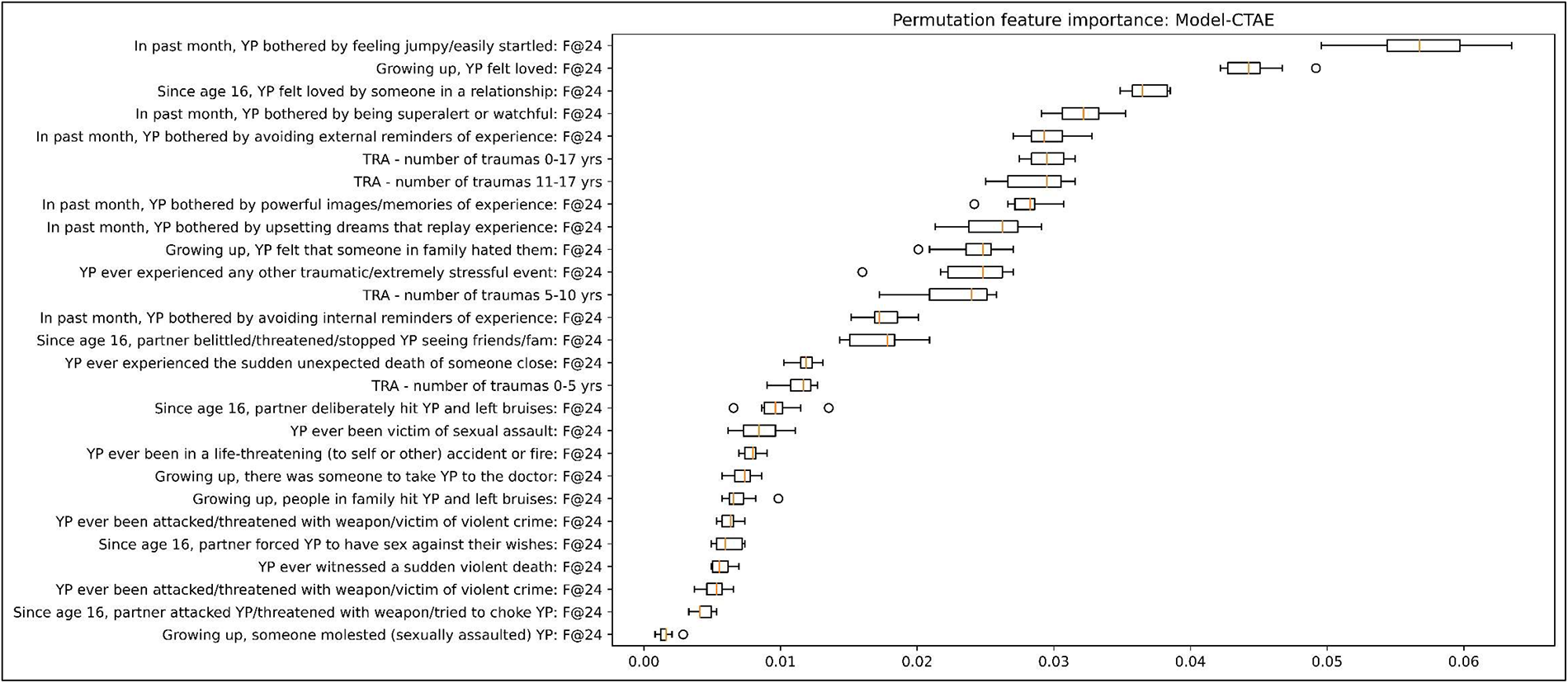
Permutation feature importance for Model-CTAE. Features ranked from most important (highest) to least important (lowest)

## Discussion

We have evaluated the performance of multivariate, life-course risk-factor data when classifying episodes of adulthood depression. Considering the prevalence of common mental health problems in the population, and recent literature calling into question the biochemical role of neurotransmitters in the aetiology of depression ^56^, approaches which focus on depression as emerging from an interplay of different factors from pre-conception (i.e., family history) to concurrent lifestyles is the most appropriate research strategy for a more person-centred understanding of depression risk. The modelling of non-linear associations between a multitude of data points related to health and wellbeing can help drive towards a broader understanding of the accumulation and combination of risk factors for depression in an exploratory manner, whilst also identifying key areas and constructs that could aid in the prevention of depression in young adults.

### Highly important trauma variables

The results of this analysis show that using information from across the entire risk ecosystem resulted in the best prediction model performance, despite only marginal differences in performance across the three highest performing models. Considering that this analysis utilised risk factor knowledge from various domains within depression research, this result is a promising sign that the feature selection process was sufficiently thorough and lays a good foundation for model development. The childhood trauma and adverse experiences feature set was the second highest performing model, supporting evidence previously highlighted that childhood trauma is a robust risk factor for adult depression ^23,24,26^. Feature importance rankings for this model showed that ‘feeling loved’ whilst growing up was amongst the top five most important features for model classification. Feeling loved as a core mental health need underscores the influence of relationship health and connectedness on future psychiatric outcomes. Valuable future work should explore how feeling loved or unloved by different caregivers, family members, and others in an individual’s support network maps onto depression outcomes, and future research should explore and prioritise relational support and relational awareness interventions as protective of mental health. Data points relating to an individual’s family history of depression not ranking high in the permutation feature importance list indicates that intergenerational family dynamics in mental health transmission may involve more than direct genetic transmission. It is worth considering however whether generational differences in ‘prevalence’ or reporting of depression may introduce some noise into the classifier. Differences in how depression has been measured in the ALSPAC resource over time is also worth considering; intergenerational depression was reported here by G0 (i.e., the parents of the core ALSPAC participants (G1)) with a single self-report question on whether their parents had a history of depression (see supplementary materials for more detail), whereas depression symptoms are measured in offspring (G1) using the SMFQ depression symptoms measurement scale.

There was also a high density of features relating to the psychological impact of trauma on the individual amongst the model’s higher performing features. Self-reported measures of trauma having a higher ability to predict than objective measures align with a recent meta-analysis showing that objective measures of childhood adversities had lower associations with future psychopathology than self-reported measures ^57^, indicating that the effects of adversity on mental health may be driven by an individual’s subjective experience. Recent research has proposed the presence of nine core features of traumatic events that align with subjective wellbeing, including constructs such as ‘impact’, ‘emotional significance’, ‘valence’, and ‘challenge’ ^58^. These constructs being positioned as central to the taxonomy of trauma supports our finding that individual differences in perceptions and judgements of trauma may be robustly linked to future mental health outcomes.

There is evidence to suggest that cumulative childhood trauma is associated with symptom complexity in adult mental health ^59^, with complex PTSD outcomes emerging from historic and sustained interpersonal traumas stemming from dysfunctional early life attachments. Individuals who experience strong psychological impact of traumatic events may be more likely to have received diagnoses of anxiety related disorders or post-traumatic stress disorder (PTSD), known to be comorbid with depression with overlapping symptomatology ^53^ . Other studies indicate however that when considering trauma as a precursor to depression, single and multiple trauma groups did not differ on indices of depressive symptoms, despite individuals with multiple trauma reporting higher levels of guilt, shame, dissociation, and interpersonal sensitivity than those who had experienced single trauma ^60^.

Within a trauma-informed framework of psychopathology ^54^, it is worth discussing whether diagnoses in these areas are more valuable than centring the individual with a symptoms-based approach that doesn’t frame reactions to highly traumatic events as ‘disordered’. Rather, psychiatric symptoms are functional and understandable responses to adverse environments and experiences, conceptualised by nondiagnostic systems such as the Power Threat Meaning Framework ^61^. Research into this area and how it aligns with clinical approaches to treatment and prevention are yet to be elucidated. Results from the model indicates that behavioural indicators of trauma are linked with depression e.g., feeling jumpy / startled, avoiding external reminders of experience, even more so than specific traumatic events e.g., being the victim of sexual or violent assault. Again, it is worth considering whether there is an underreporting of explicit violence and abuse in survey data due to a variety of reasons, such as common issues around retrospective recall and memory issues when reporting trauma. It could also be possible that those survey respondents with the highest degree of trauma may not participate in self-report methodologies of this sort due to finding the question content triggering, leading to further missingness within the data. Taking the results at face value, there is an indication that individual psychological illness burden is predictive of depression, rather than experience of trauma outright. Whether this effect is mediated by individual resilience requires further research and indices of resilience to be included as a feature within the model, with mechanisms of individual resilience having been proposed ^62^ but not yet fully understood. Emotional resilience has been suggested as mediating ‘post-traumatic growth’ following adverse childhood experiences ^63^ alongside event centrality, a measure of how much adverse experiences significantly alter the trajectory of an individual’s life. The results of this modelling approach which combines indices of traumatic events in addition to measurements of trauma impact indicates that exposure to adversity alone may not be a robust, individual predictor, and is unable to differentially explain psychopathological outcomes in the absence of other data relating to subjective experiences of the impact and significance of trauma.

These results suggest that improving measurement and prevention of early childhood trauma and adverse experiences should continue to be prioritised in mental health interventions. There is ongoing work within public health authorities in the United Kingdom (UK) ^64^ to intervene on and prevent adverse childhood events (ACEs) occurring in childhood, through efforts to disseminate information and share evidence that will help mitigate the effects of ACEs in families and communities.

### Predictive power of sleep health and self-appraisal of wellbeing

Another important feature in the best performing model were problems getting to sleep at age 24, in line with the view that sleep operates as both a risk factor for and symptom of both depression and anxiety ^35^. Circadian rhythms are well evidenced as playing an aetiological role in mental illness, with significant amounts of research funding ^65^ going towards prioritising sleep research as a core determinant and preventative target for the management of these conditions. Another consideration to make in light of this result is that some antidepressant agents are known to have a disruptive effect on sleep quality ^66^, which could introduce complexities relating to directionality and causal inference in this area if a certain number of participants in the sample have managed their depression symptoms with medication. The addition of medication data into these models would provide useful insight into the relationship between pharmacological intervention for depression and sleep, and facilitate further investigation into this finding. Considering this result in tandem with self-perception of health having high importance in both Model-AF and Model-HBL provide evidence that subjective perception of health is a reliable indicator of objective symptom manifestations. This is in line with previous research which has found associations between early adolescent depression and poorer self-perceived general health ^6,7^.This insight should be interpreted as encouraging that self-reported data points that are easy and concise to measure and implement have predictive utility in the absence of other screening data.

### Changes in model accuracy after permuting features

When extrapolating the results of permuted feature importance rankings to the broader epidemiological context, it is important to note that the ranking of each feature within a particular model serves a determination of how useful that feature was for classification within that model only and may be differently ranked in a model containing different features or training data. There was also variability in the degree to which accuracy changed when permuting features within each individual model, with low changes in accuracy making interpretation of the overall hierarchy of importance more challenging. For more information, consult the sci-kit learn permutation feature importance software documentation ^67^.

A general consideration for the model performances is that all models underperformed when classifying individuals in the ‘depressed’ category compared to ‘non-depressed’. This could either indicate that not being depressed is easier to predict from the risk factors included across all 4 models, or that there is heterogeneity in the pool of depressed participants which make their outcomes harder to predict. A potential explanation is also that positive cases across the risk factors (i.e., presence of maltreatment in childhood, physical comorbidities) are relatively rare compared to negative cases, so the models may be biased towards classifying people as not depressed.

### Conclusion and limitations

This paper has explored a variety of factors related to early adulthood depression, and has measured the predictive ability of these factors when aggregated into individual conceptual silos. We have shown that depression can be classified using a range of factors measuring various health related constructs in a birth cohort population. The model which utilised the entire breadth of risk factor data related to depression performed the best, emphasising the utility of high-throughput approaches to depression prediction spanning multiple domains of risk. There are a few considerations and limitations to this approach which will be outlined below.

The domains, features, and stratifications included in this paper are not functioning as an exhaustive list of every potential or relevant risk factor for depression in young adults. The conceptualisation of each individual risk domain could be improved with the addition of more information from a much wider array of factors relating to not only depression risk but depression prevention and management. Rather than aiming to conceptually model the entire risk ecosystem of depression in its entirety, this work serves as a starting point for how to integrate longitudinal measures of depression risk into a ML based prediction model that is highly interpretable. Stratifying risk factors into specific domains is not a perfect process, with many factors having behavioural, psychological, endocrine, genetic, and molecular substrates. An important consideration for risk factor stratification is that factors are interconnected and complex, and exist as a network, rather than independent silos. Despite this, measurement of specific domains gives insight into which sort of information to prioritise when building mental health prediction models from longitudinal screening data.

Model-AF was limited due to exclusion of the PRS for depression variable. This variable was excluded due to the use of imputation of missing data in that model, due to imputation of a genetic variable reducing analysis accuracy.

The ALSPAC study is a multigenerational, longitudinal data resource spanning multiple decades. Participants are free to engage as little or as often as they like with questionnaires, clinic visits, and other data collection activities whilst remaining in the study, and are also free to withdraw consent for their data to be used at any time. For this reason, management of missing data is an ongoing challenge and consideration when building and training high-throughput, data driven models. For more information on how missing data was handled in this analysis, please see the methods section.

In conclusion, this study has utilised a broad variety of healthcare data to predict a longitudinal depression symptom variable derived from measuring depression at multiple timepoints, mitigating the risk of measurement error when utilising data points collected at single timepoints, as well as incorporating the episodic and remising nature of depression into the analysis design. Use of longitudinal data here is a key strength of this modelling approach which future work using this resource can expand on.

## Methods

### Setting

The Avon Longitudinal Study of Parents and Children (ALSPAC) is a population birth cohort study launched in the 1990s that followed the lives of approximately 15,000 families in Bristol and the surrounding areas ^68,69^. Pregnant women resident in Avon, UK with expected dates of delivery between 1st April 1991 and 31st December 1992 were invited to take part in the study. 20,248 pregnancies have been identified as being eligible and the initial number of pregnancies enrolled was 14,541. Of the initial pregnancies, there was a total of 14,676 foetuses, resulting in 14,062 live births and 13,988 children who were alive at 1 year of age. When the oldest children were approximately 7 years of age, an attempt was made to bolster the initial sample with eligible cases who had failed to join the study originally. As a result, when considering variables collected from the age of seven onwards (and potentially abstracted from obstetric notes) there are data available for more than the 14,541 pregnancies mentioned above: The number of new pregnancies not in the initial sample (known as Phase I enrolment) that are currently represented in the released data and reflecting enrolment status at the age of 24 is 906, resulting in an additional 913 children being enrolled (456, 262 and 195 recruited during Phases II, III and IV respectively). The total sample size for analyses using any data collected after the age of seven is therefore 15,447 pregnancies, resulting in 15,658 foetuses. Of these 14,901 children were alive at 1 year of age ^70^.

The types of data available in ALSPAC is vast and spans many different domains of health, lifestyle, and wellbeing. Data have been collected across the ALSPAC resource using a variety of self-report, in-clinic, and biological assays, with online data being captured through REDcap software ^71^. Details about ALSPAC, an overview of available data, and a fully searchable data dictionary and variable search tool can be found at the following webpage: http://www.bristol.ac.uk/alspac/researchers/our-data/. Study data were collected and managed using REDCap electronic data capture tools hosted at the University of Bristol. REDCap (Research Electronic Data Capture) is a secure, web-based software platform designed to support data capture for research studies ^72^. Ethical approval for the study was obtained from the ALSPAC Ethics and Law Committee and the Local Research Ethics Committees. Informed consent for the use of data collected via questionnaires and clinics was obtained from participants following the recommendations of the ALSPAC Ethics and Law Committee at the time.

### Outcomes and predictors

Depression is defined in the context of this analysis as participants that have experienced at least one episode of depression between the ages of 23 and 28. An episode is defined as having a score of >=11 on the Short Mood and Feelings Questionnaire (SMFQ) ^50^, a commonly used depression screening tool^73^.

We examined a number of predictors drawn from across the life course. A full list of these predictors can be found in Table 1 in the main text and table 1 in the supplementary materials.

### Data cleaning

Data were extracted (n=15,645) from the ALSPAC repository using STATA Software Version 16. Variables included a mixture of both raw measurements and derived variables. Participants that did not have a value for the model target variable (any episodes of depression post-24 years old) were removed (n=5,731).

### Handling missingness

All values used to indicate different levels of data missingness (e.g., values to indicate a participant not showing up to a clinic; participants not completing a questionnaire; missing individual questionnaire items) were set to NaN. Individual features and rows that had >=50% NaN values were dropped (n=3,487.). The rest of the missing data (excluding the target variable) were imputed with a K Nearest Neighbours (KNN) imputation algorithm, using the Scikit Learn Python package (Python version 3.8.13). KNN imputation was utilised to reduce missingness within the data due to the ability of the algorithm to handle a variety of different data types, and its lack of assumptions about linearity of data.

### Random Forest Model

The Random Forest algorithm was implemented using scikit-learn and Python version 3.8.13. RandomizedSearch cross validation (CV) approach with 5 folds was used to select optimal parameters for the model. Tuned hyperparameters and their ranges can be found in Table 4 below. Each feature set underwent individual iterations of hyperparameter tuning to create bespoke parameters for each model.

**Table 4:**
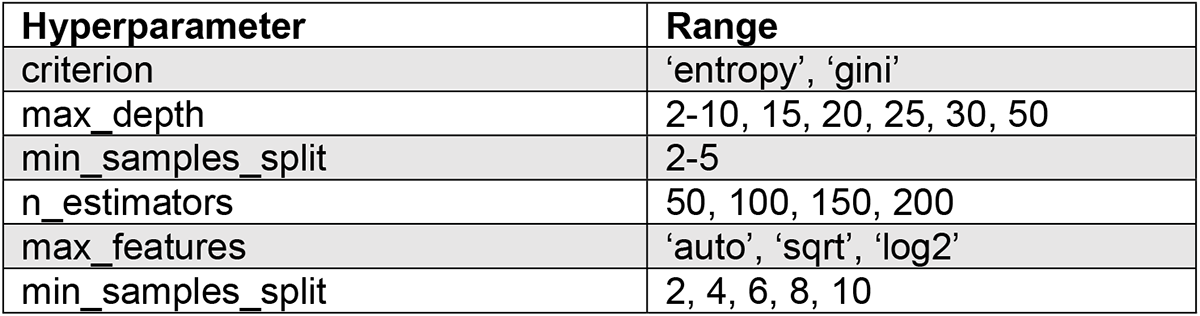
Hyperparameters and ranges searched over to find optimal parameters across all models.

| Hyperparameter | Range |
| --- | --- |
| criterion | 'entropy', 'gini' |
| max_depth | 2-10, 15, 20, 25, 30, 50 |
| min_samples_split | 2-5 |
| n_estimators | 50, 100, 150, 200 |
| max_features | 'auto', 'sqrt', 'log2' |
| min_samples_split | 2, 4, 6, 8, 10 |

### Permutation feature importance

The permutation feature importance ranking protocol was based on scikit learn documentation for use with multicollinear features ^74^. Changes in model accuracy after features are permuted from the train data set are used to determine how much the model relies on each feature during model training.

### Code availability statement

The underlying code for this study is available in ‘Modelling the risk ecosystem of depression_randomforest_code.’ and can be accessed via this link https://osf.io/3k2d9/?view_only=2ebc272b4c02464e8eb20688bbfea929.

## Data Availability

ALSPAC data access is through a system of managed open access. The steps below highlight how to apply for access to ALSPAC data:

1. Please read the ALSPAC access policy which describes the process of accessing the data in detail, and outlines the costs associated with doing so.
2. You may also find it useful to browse the fully searchable research proposals database, which lists all research projects that have been approved since April 2011.
3. Please submit your research proposal for consideration by the ALSPAC Executive Committee. You will receive a response within 10 working days to advise you whether your proposal has been approved. If you have any questions about accessing data, please email.

## Acknowledgements

We are extremely grateful to all the families who took part in this study, the midwives for their help in recruiting them, and the whole ALSPAC team, which includes interviewers, computer and laboratory technicians, clerical workers, research scientists, volunteers, managers, receptionists, and nurses.

## Authorship contributions

Conceptualization: HF, ASFK, BID, RM, RP

Methodology: HF, ASFK, BID, RM, RP

Formal Analysis: HF, ASFK

Data Curation: HF, ASFK

Writing Original Draft: HF

Writing Review & Editing: HF, ASFK, MB, BID, RM, RP

Supervision: ASFK, BID, RM, RP

All authors read and approved the final manuscript.

## References

1. Kessler, R. C. et al. Age of onset of mental disorders: A review of recent literature. Curr Opin Psychiatry 20, 359–364 (2007).

2. Kwong, A. S. F. Examining the longitudinal nature of depressive symptoms in the Avon Longitudinal Study of Parents and Children (ALSPAC). Wellcome Open Res 4, (2019).

3. Pedrelli, P., Shapero, B., Archibald, A. & Dale, C. Alcohol use and depression during adolescence and young adulthood: a summary and interpretation of mixed findings. Curr Addict Rep 3, 91–97 (2016).

4. Solmi, M. et al. Age at onset of mental disorders worldwide: large-scale meta-analysis of 192 epidemiological studies. Mol Psychiatry 27, 281–295 (2022).

5. Johnson, D., Dupuis, G., Piche, J., Clayborne, Z. & Colman, I. Adult mental health outcomes of adolescent depression: A systematic review. Depression and Anxiety 35, 700–716 (2018).

6. Miller, D. K., Constance, H. L. & Brennan, P. A. Health Outcomes Related to Early Adolescent Depression. J Adolesc Health 41, 256–262 (2007).

7. Naicker, K., Galambos, N. L., Zeng, Y., Senthilselvan, A. & Colman, I. Social, Demographic, and Health Outcomes in the 10 Years Following Adolescent Depression. Journal of Adolescent Health 52, 533–538 (2013).

8. Kwong, A. S. F. et al. Genetic and Environmental Risk Factors Associated With Trajectories of Depression Symptoms From Adolescence to Young Adulthood. JAMA Network Open 2, e196587 (2019).

9. Albert, P. R. Why is depression more prevalent in women? J Psychiatry Neurosci 40, 219–221 (2015).

10. Kuehner, C. Why is depression more common among women than among men? The Lancet Psychiatry 4, 146–158 (2017).

11. Salk, R. H., Hyde, J. S. & Abramson, L. Y. Gender differences in depression in representative national samples: Meta-analyses of diagnoses and symptoms. Psychol Bull 143, 783–822 (2017).

12. Howard, D. M. et al. Genome-wide meta-analysis of depression identifies 102 independent variants and highlights the importance of the prefrontal brain regions. Nat Neurosci 22, 343–352 (2019).

13. Howard, D. M. et al. Genome-wide association study of depression phenotypes in UK Biobank identifies variants in excitatory synaptic pathways. Nat Commun 9, 1470 (2018).

14. Levey, D. F. et al. Bi-ancestral depression GWAS in the Million Veteran Program and meta-analysis in >1.2 million individuals highlight new therapeutic directions. Nat Neurosci 24, 954–963 (2021).

15. Wray, N. R. et al. Genome-wide association analyses identify 44 risk variants and refine the genetic architecture of major depression. Nat Genet 50, 668–681 (2018).

16. Cao, Z. et al. Polygenic risk score, healthy lifestyles, and risk of incident depression. Transl Psychiatry 11, 1–9 (2021).

17. Halldorsdottir, T. et al. Polygenic Risk: Predicting Depression Outcomes in Clinical and Epidemiological Cohorts of Youths. Am J Psychiatry 176, 615–625 (2019).

18. Stein, A. et al. Effects of perinatal mental disorders on the fetus and child. The Lancet 384, 1800–1819 (2014).

19. Halonen, J., Hakko, H., Riala, K. & Riipinen, P. Familial Risk Factors in Relation to Recurrent Depression Among Former Adolescent Psychiatric Inpatients. Child Psychiatry Hum Dev 53, 515–525 (2022).

20. Rasic, D., Hajek, T., Alda, M. & Uher, R. Risk of Mental Illness in Offspring of Parents With Schizophrenia, Bipolar Disorder, and Major Depressive Disorder: A Meta-Analysis of Family High-Risk Studies. Schizophr Bull 40, 28–38 (2014).

21. Houtepen, L. C., Heron, J., Suderman, M. J., Tilling, K. & Howe, L. D. Adverse childhood experiences in the children of the Avon Longitudinal Study of Parents and Children (ALSPAC). Wellcome Open Res 3, 106 (2018).

22. Dunn, E. C., Nishimi, K., Powers, A. & Bradley, B. Is developmental timing of trauma exposure associated with depressive and post-traumatic stress disorder symptoms in adulthood? J Psychiatr Res 84, 119–127 (2017).

23. Negele, A., Kaufhold, J., Kallenbach, L. & Leuzinger-Bohleber, M. Childhood Trauma and Its Relation to Chronic Depression in Adulthood. Depress Res Treat 2015, 650804 (2015).

24. McLaughlin, K. A. et al. Trauma exposure and posttraumatic stress disorder in a national sample of adolescents. J Am Acad Child Adolesc Psychiatry 52, 815–830.e14 (2013).

25. Lewis, S. J. et al. The epidemiology of trauma and post-traumatic stress disorder in a representative cohort of young people in England and Wales. The Lancet Psychiatry 6, 247–256 (2019).

26. Schalinski, I. et al. Type and timing of adverse childhood experiences differentially affect severity of PTSD, dissociative and depressive symptoms in adult inpatients. BMC Psychiatry 16, 295 (2016).

27. Vaughn-Coaxum, R. A., Wang, Y., Kiely, J., Weisz, J. R. & Dunn, E. C. Associations Between Trauma Type, Timing, and Accumulation on Current Coping Behaviors in Adolescents: Results from a Large, Population-based Sample. J Youth Adolesc 47, 842– 858 (2018).

28. Ahuja, M., Okoro, J., Frimpong, E., Doshi, R. P. & Wani, R. J. Feeling Not Wanted/Loved and Depression: Does Gender Matter? Psychol Rep 00332941211062822 (2021) doi:10.1177/00332941211062822.

29. Brunstein klomek, A., Marrocco, F., Kleinman, M., Schonfeld, I. S. & Gould, M. S. Bullying, Depression, and Suicidality in Adolescents. Journal of the American Academy of Child & Adolescent Psychiatry 46, 40–49 (2007).

30. Kaltiala-Heino, R., Fröjd, S. & Marttunen, M. Involvement in bullying and depression in a 2-year follow-up in middle adolescence. Eur Child Adolesc Psychiatry 19, 45–55 (2010).

31. Goodman, R. J., Samek, D. R., Wilson, S., Iacono, W. G. & McGue, M. Close relationships and depression: A developmental cascade approach. Dev Psychopathol 31, 1451–1465 (2019).

32. Steffen, A., Nübel, J., Jacobi, F., Bätzing, J. & Holstiege, J. Mental and somatic comorbidity of depression: a comprehensive cross-sectional analysis of 202 diagnosis groups using German nationwide ambulatory claims data. BMC Psychiatry 20, 142 (2020).

33. Noh, J.-W., Kwon, Y. D., Park, J., Oh, I.-H. & Kim, J. Relationship between Physical Disability and Depression by Gender: A Panel Regression Model. PLoS One 11, e0166238 (2016).

34. Shen, S.-C., Huang, K.-H., Kung, P.-T., Chiu, L.-T. & Tsai, W.-C. Incidence, risk, and associated factors of depression in adults with physical and sensory disabilities: A nationwide population-based study. PLoS One 12, e0175141 (2017).

35. Nutt, D., Wilson, S. & Paterson, L. Sleep disorders as core symptoms of depression. Dialogues Clin Neurosci 10, 329–336 (2008).

36. Bridges, S. & Disney, R. Debt and depression. Journal of Health Economics 29, 388–403 (2010).

37. Butterworth, P., Rodgers, B. & Windsor, T. D. Financial hardship, socio-economic position and depression: Results from the PATH Through Life Survey. Social Science & Medicine 69, 229–237 (2009).

38. Guan, N., Guariglia, A., Moore, P., Xu, F. & Al-Janabi, H. Financial stress and depression in adults: A systematic review. PLOS ONE 17, e0264041 (2022).

39. Skapinakis, P., Weich, S., Lewis, G., Singleton, N. & Araya, R. Socio-economic position and common mental disorders: Longitudinal study in the general population in the UK. The British Journal of Psychiatry 189, 109–117 (2006).

40. Sweet, E., Nandi, A., Adam, E. K. & McDade, T. W. The high price of debt: Household financial debt and its impact on mental and physical health. Social Science & Medicine 91, 94–100 (2013).

41. Sebastian, C. L., Pote, I. & Wolpert, M. Searching for active ingredients to combat youth anxiety and depression. Nat Hum Behav 5, 1266–1268 (2021).

42. Edwards, A. C. et al. Adolescent Alcohol Use Is Positively Associated With Later Depression in a Population-Based U.K. Cohort. J Stud Alcohol Drugs 75, 758–765 (2014).

43. Kuria, M. W. et al. The Association between Alcohol Dependence and Depression before and after Treatment for Alcohol Dependence. ISRN Psychiatry 2012, 482802 (2012).

44. Mason, M., Mennis, J., Russell, M., Moore, M. & Brown, A. Adolescent Depression and Substance Use: The Protective Role of Prosocial Peer Behavior. J Abnorm Child Psychol 47, 1065–1074 (2019).

45. Wang, P.-W. & Yen, C.-F. Adolescent substance use behavior and suicidal behavior for boys and girls: a cross-sectional study by latent analysis approach. BMC Psychiatry 17, 392 (2017).

46. Chen, C.-Y., Wagner, F. A. & Anthony, J. C. Marijuana use and the risk of Major Depressive Episode. Epidemiological evidence from the United States National Comorbidity Survey. Soc Psychiatry Psychiatr Epidemiol 37, 199–206 (2002).

47. Degenhardt, L., Hall, W. & Lynskey, M. Exploring the association between cannabis use and depression. Addiction 98, 1493–1504 (2003).

48. Degenhardt, L. et al. The persistence of the association between adolescent cannabis use and common mental disorders into young adulthood. Addiction 108, 124–133 (2013).

49. Prediction of the trajectories of depressive symptoms among children in the adolescent brain cognitive development (ABCD) study using machine learning approach. Journal of Affective Disorders 310, 162–171 (2022).

50. Turner, N., Joinson, C., Peters, T. J., Wiles, N. & Lewis, G. Validity of the Short Mood and Feelings Questionnaire in late adolescence. Psychological Assessment 26, 752–762 (2014).

51. sklearn.dummy.DummyClassifier. *scikit-learn* https://scikit-learn/stable/modules/generated/sklearn.dummy.DummyClassifier.html.

52. Molnar, C. *8.5 Permutation Feature Importance | Interpretable Machine Learning*.

53. Flory, J. D. & Yehuda, R. Comorbidity between post-traumatic stress disorder and major depressive disorder: alternative explanations and treatment considerations. Dialogues Clin Neurosci 17, 141–150 (2015).

54. Sweeney, A., Filson, B., Kennedy, A., Collinson, L. & Gillard, S. A paradigm shift: relationships in trauma-informed mental health services. BJPsych Adv 24, 319–333 (2018).

55. Kalin, N. H. The Critical Relationship Between Anxiety and Depression. AJP 177, 365– 367 (2020).

56. Moncrieff, J. et al. The serotonin theory of depression: a systematic umbrella review of the evidence. Mol Psychiatry 1–14 (2022) doi:10.1038/s41380-022-01661-0.

57. Francis, E. R., Tsaligopoulou, A., Stock, S. E., Pingault, J.-B. & Baldwin, J. R. Subjective and objective experiences of childhood adversity: a meta-analysis of their agreement and relationships with psychopathology. Journal of Child Psychology and Psychiatry **n/a**,.

58. Luhmann, M., Fassbender, I., Alcock, M. & Haehner, P. A dimensional taxonomy of perceived characteristics of major life events. Journal of Personality and Social Psychology 121, 633–668 (2021).

59. Cloitre, M. et al. A developmental approach to complex PTSD: Childhood and adult cumulative trauma as predictors of symptom complexity. Journal of Traumatic Stress 22, 399–408 (2009).

60. Hagenaars, M. A., Fisch, I. & van Minnen, A. The effect of trauma onset and frequency on PTSD-associated symptoms. Journal of Affective Disorders 132, 192–199 (2011).

61. Johnstone, L. & Boyle, M. The Power Threat Meaning Framework: An Alternative Nondiagnostic Conceptual System. Journal of Humanistic Psychology 0022167818793289 (2018) doi:10.1177/0022167818793289.

62. Yi, F., Li, X., Song, X. & Zhu, L. The Underlying Mechanisms of Psychological Resilience on Emotional Experience: Attention-Bias or Emotion Disengagement. Front Psychol 11, 1993 (2020).

63. Tranter, H., Brooks, M. & Khan, R. Emotional resilience and event centrality mediate posttraumatic growth following adverse childhood experiences. *Psychological Trauma: Theory, Research*, Practice, and Policy 13, 165–173 (2021).

64. Adverse Childhood Experiences. *Public Health Wales* https://phw.nhs.wales/topics/adverse-childhood-experiences/.

65. Sleep science and mental health | Grant funding. *Wellcome* https://wellcome.org/grant-funding/schemes/mental-health-award-integrating-sleep-and-circadian-science-our-understanding.

66. Wichniak, A., Wierzbicka, A., Walęcka, M. & Jernajczyk, W. Effects of Antidepressants on Sleep. Curr Psychiatry Rep 19, 63 (2017).

67. 4.2. Permutation feature importance. *scikit-learn* https://scikit-learn/stable/modules/permutation_importance.html.

68. Boyd, A. et al. Cohort Profile: The ‘Children of the 90s’—the index offspring of the Avon Longitudinal Study of Parents and Children. International Journal of Epidemiology 42, 111–127 (2013).

69. Fraser, A. et al. Cohort Profile: the Avon Longitudinal Study of Parents and Children: ALSPAC mothers cohort. Int J Epidemiol 42, 97–110 (2013).

70. Northstone, K. et al. The Avon Longitudinal Study of Parents and Children (ALSPAC): an update on the enrolled sample of index children in 2019. Wellcome Open Res 4, 51 (2019).

71. REDCap. https://projectredcap.org/.

72. Harris, P. A. et al. Research electronic data capture (REDCap)--a metadata-driven methodology and workflow process for providing translational research informatics support. J Biomed Inform 42, 377–381 (2009).

73. Angold, A., Costello, E. J., Messer, S. C. & Pickles, A. Development of a short questionnaire for use in epidemiological studies of depression in children and adolescents. International Journal of Methods in Psychiatric Research 5, 237–249 (1995).

74. Permutation Importance with Multicollinear or Correlated Features. *scikit-learn* https://scikit-learn/stable/auto_examples/inspection/plot_permutation_importance_multicollinear.html.

75. Croft, J. et al. Association of Trauma Type, Age of Exposure, and Frequency in Childhood and Adolescence With Psychotic Experiences in Early Adulthood. JAMA Psychiatry 76, 79–86 (2019).

76. Psychometric Properties of the Life Events Checklist – Matt J. Gray, Brett T. Litz, Julie L. Hsu, Thomas W. Lombardo, 2004. https://journals.sagepub.com/doi/abs/10.1177/1073191104269954?casa_token=4WFHNXC8lMMAAAAA:hEDZJxi2WDrGGn3GES4J2zjzBhQYvGKrcxYyrnwNbrKRbnDN2AmpZKWYTYM-GhzmTyrbB14cW0s.

77. Choi, S. W. & O’Reilly, P. F. PRSice-2: Polygenic Risk Score software for biobank-scale data. GigaScience 8, giz082 (2019).

78. Kwong, A. S. F. et al. Polygenic risk for depression, anxiety and neuroticism are associated with the severity and rate of change in depressive symptoms across adolescence. Journal of Child Psychology and Psychiatry 62, 1462–1474 (2021).

79. Piontek, D., Kraus, L., Legleye, S. & Bühringer, G. The validity of DSM-IV cannabis abuse and dependence criteria in adolescents and the value of additional cannabis use indicators. Addiction 106, 1137–1145 (2011).

