## Supplementary materials for "Modelling the risk ecosystem of depression using machine learning in a population of young adults"

*Detailed table of exposures*

| Exposure description | ALSPAC variable name | Informant | Response | Assessed | Question / item |
|---|---|---|---|---|---|
| Sex | kz021 | Mother | Male/female | Questionnaires | NA |
| Family history of depression and mental health difficulties<br>Mother's 'About yourself' questionnaire, Section A 'Your medical history' and father's 'Your environment' questionnaire, Section D 'Your medical history' | | | | | |
| Mother had severe depression | d171a | Mother | Yes (1) / No (2) | 12weeks gestation, questionnaire | QA9: 'Have you ever had severe depression?' |
| Mother had any other psychiatric problem | d172a | Mother | Yes (1) / No (2) | 12weeks gestation, questionnaire | QA9: 'Have you ever had any other psychiatric problem?' |
| Maternal grandmother had depression or 'nerves' | d536 | Mother | Yes (1) / No (2) / Don't know (9) | 12weeks gestation, questionnaire | QC9: 'Has your natural mother and/or mother figure had… depression or 'nerves'?' (Mother 'About Yourself' questionnaire; Section C 'You and Your Parents') |
| Maternal grandfather had depression or 'nerves' | d586 | Mother | Yes (1) / No (2) | 12 weeks gestation, questionnaire | QC12: 'Has your natural father and/or father figure had… depression or 'nerves'?' (Mother 'About Yourself' questionnaire; Section C 'You and Your Parents') |
| Partner had severe depression | pa191a | Partner | Yes (1) / No (2) | 12weeks gestation, questionnaire | QD6: 'Have you ever had severe depression?' (Father 'Your environment' questionnaire, Section D 'Your Medical History') |

| | | | | | |
|---|---|---|---|---|---|
| | | | | (and 18 weeks gestation) | |
| Partner had any other psychiatric problem | pa192a | Partner | Yes (1) / No (2) | 12weeks gestation, questionnaire and 18 weeks gestation | QD6: 'Have you ever had any other psychiatric problem?' (Father 'Your Environment' questionnaire, Section D 'Your Medical History') |
| Paternal grandmother had depression or 'nerves' | pa536 | Partner | Yes (1) / No (2) / Don't know (9) | 12 weeks gestation, questionnaire | QF9: 'Has your natural mother and/or mother figure had… depression or 'nerves'' (Father 'Your Environment' questionnaire; Section F 'You and Your Parents' |
| Paternal grandfather had depression or 'nerves' | pa586 | Partner | Yes (1) / No (2) / Don't know (9) | 12 weeks gestation, questionnaire | QF12: 'Has your natural father and/or father figure had… depression or 'nerves'?' (Father 'Your Environment' questionnaire; Section F 'You and Your Parents' |
| <u>Traumatic childhood experiences</u><br>Trauma measures were derived from a mixture of parent and child self-report, from 48 separate assessments from ages 0-17. Data were also supplemented by child self-report at age 22. For more detail see Croft et al. [75] for information on how trauma variables were assessed and derived | | | | | |
| Number of traumas from 0-17 years | clon170 | Parent and child | 0-6 | Collected prospectively from ages 0-17 and supplemented by retrospective data at age 22 | See Croft et al. [75] for information on how trauma variables were assessed and derived |
| Number of traumas from 0-5 years | clon167 | | 0-4 | | |
| Number of traumas from 5-10 years | clon168 | | 0-6 | | |
| Number of traumas from 11-17 years | clon169 | | 0-4 | | |
| <u>Difficult experiences questionnaire</u><br>Traumatic experiences and symptoms of post-traumatic stress disorder (PTSD) | | | | | |

| | | | | | |
|---|---|---|---|---|---|
| Assessed at the 'Focus @ 24' clinic at age 24<br>The traumatic experiences questions include questions on childhood abuse and neglect, as used in the UK E-Risk child/adolescent cohort and in UK Biobank<br><br>Questions on relationship abuse (since the age of 16) are based on questions used in the British Crime Survey on Domestic Abuse.<br><br>Questions on life-time exposure to traumatic events are from the Life Events Checklist for DSM-V (Gray et al., 2004) [76]. | | | | | |
| Feeling loved in childhood | FKDE1010 | Child | Never true (1) / Rarely true (2) / Sometimes true (3) / Often true (4) / Very often true (5) | 24 years, clinic questionnaire | 'Growing up, young person (YP) felt loved' |
| Physical abuse in childhood | FKDE1020 | Child | Never true (1) / Rarely true (2) / Sometimes true (3) / Often true (4) / Very often true (5) | 24 years, clinic questionnaire | 'Growing up, people in family hit YP and left bruises' |
| Feeling hated in childhood | FKDE1030 | Child | Never true (1) / Rarely true (2) / Sometimes true (3) / Often true (4) / Very often true (5) | 24 years, clinic questionnaire | 'Growing up, YP felt that someone in family hated them' |
| Sexual abuse in childhood | FKDE1040 | Child | Never true (1) / Rarely true (2) / Sometimes true (3) / Often true (4) / Very often true (5) | 24 years, clinic questionnaire | 'Growing up, someone molested (sexually assaulted) YP' |
| Access to medical care | FKDE1050 | Child | Never true (1) / Rarely true (2) / Sometimes true (3) / Often true | 24 years, clinic questionnaire | 'Growing up, there was someone to take YP to the doctor' |

| | | | | | |
|---|---|---|---|---|---|
| | | | (4) / Very often true (5) | | |
| Feeling loved by someone in a relationship in adolescence | FKDE1110 | Child | Never true (1) / Rarely true (2) / Sometimes true (3) / Often true (4) / Very often true (5) | 24 years, clinic questionnaire | ‘Since age 16, YP felt loved by someone in a relationship’ |
| Domestic abuse in romantic relationship in adolescence | FKDE1120 | Child | Never true (1) / Rarely true (2) / Sometimes true (3) / Often true (4) / Very often true (5) | 24 years, clinic questionnaire | ‘Since age 16, partner deliberately hit YP and left bruises’ |
| Domestic abuse in romantic relationship in adolescence | FKDE1130 | Child | Never true (1) / Rarely true (2) / Sometimes true (3) / Often true (4) / Very often true (5) | 24 years, clinic questionnaire | ‘Since age 16, partner attacked YP/threatened with weapon/tried to choke YP’ |
| Domestic abuse in romantic relationship in adolescence | FKDE1140 | Child | Never true (1) / Rarely true (2) / Sometimes true (3) / Often true (4) / Very often true (5) | 24 years, clinic questionnaire | ‘Since age 16, partner belittled/threatened/stopped YP seeing friends/family’ |
| Sexual abuse in romantic relationship in adolescence | FKDE1150 | Child | Never true (1) / Rarely true (2) / Sometimes true (3) / Often true (4) / Very often true (5) | 24 years, clinic questionnaire | ‘Since age 16, partner forced YP to have sex against their wishes’ |
| Been in life-threatening accident or fire | FKDE1210 | Child | Yes (1) / No (0) | 24 years, clinic questionnaire | ‘YP ever been in a life-threatening (so self or other) accident or fire’ |

| | | | | | |
|---|---|---|---|---|---|
| Been a victim of violent crime | FKDE1220 | Child | Yes (1) / No (0) | 24 years, clinic questionnaire | 'YP ever been attacked/threatened with weapon/victim of violent crime' |
| Been a victim of sexual assault | FKDE1230 | Child | Yes (1) / No (0) | 24 years, clinic questionnaire | 'YP ever been victim of sexual assault' |
| Witnessed a sudden violent death | FKDE1240 | Child | Yes (1) / No (0) | 24 years, clinic questionnaire | 'YP ever witnessed a sudden violent death' |
| Experienced sudden death of someone close | FKDE1250 | Child | Yes (1) / No (0) | 24 years, clinic questionnaire | 'YP ever experienced the sudden unexpected death of someone close' |
| Experienced other traumatic / stressful event | FKDE1260 | Child | Yes (1) / No (0) | 24 years, clinic questionnaire | 'YP ever experienced any other traumatic/extremely stressful event' |
| Experienced upsetting dreams that replay experience | FKDE1310 | Child | Not at all (0), A little bit (1), Moderately (2), Quite a bit (3), Extremely (4) | 24 years, clinic questionnaire | 'In past month, YP bothered by upsetting dreams that replay experience' |
| Experienced flashbacks related to traumatic experience | FKDE1320 | Child | Not at all (0), A little bit (1), Moderately (2), Quite a bit (3), Extremely (4) | 24 years, clinic questionnaire | 'In past month, YP bothered by powerful images/memories of experience' |
| Experienced flashbacks related to traumatic experience | FKDE1330 | Child | Not at all (0), A little bit (1), Moderately (2), Quite a bit (3), Extremely (4) | 24 years, clinic questionnaire | 'In past month, YP bothered by avoiding internal reminders of experience' |
| Experienced flashbacks related to traumatic experience | FKDE1340 | Child | Not at all (0), A little bit (1), Moderately (2), Quite a bit (3), Extremely (4) | 24 years, clinic questionnaire | 'In past month, YP bothered by avoiding external reminders of experience' |
| Experienced hypervigilance | FKDE1350 | Child | Not at all (0), A little bit (1), | 24 years, clinic questionnaire | 'In past month, YP bothered by being "superalert" or watchful' |

| | | | | | |
|---|---|---|---|---|---|
| | | | Moderately (2), Quite a bit (3), Extremely (4) | | |
| Experienced hypervigilance | FKDE1360 | Child | Not at all (0), A little bit (1), Moderately (2), Quite a bit (3), Extremely (4) | 24 years, clinic questionnaire | 'In past month, YP bothered by feeling jumpy/easily startled' |
| <u>Health and wellbeing</u><br>Measures of health, illness and general wellbeing measured from the 'Your Life Now' questionnaire assessed at age 21, Life at 22+ questionnaire, and 'Focus@24' clinic at age 24 | | | | | |
| Self-assessment of health | YPA4000 | Child | Excellent (1), Very good (2), Good (3), Fair (4), Poor (5) | 21+ years, questionnaire | QD1: 'In general, YP would say their health is…' |
| Self-assessment of health | YPB1000 | Child | Excellent (1), Very good (2), Good (3), Fair (4), Poor (5) | 22+ years, questionnaire | QA1: 'Respondent's assessment of general health' |
| Has long-term illness, disability or infirmity | FKDQ2520 | Child | Yes (1), No (2) | 24 years, clinic questionnaire | 'YP has a long-standing illness, disability or infirmity' |
| <u>Polygenic risk score for depression</u><br>The MDD PRS was created using PRSice-2 [77] using summary statistics from a recent genome-wide association study (GWAS) of depression (aka broad depression [12]). ALSPAC was not included in this GWAS. The PRS was created by weighting the effect sizes of the single-nucleotide polymorphisms (SNPs) associated with depression from the initial GWAS at nine *p*-value thresholds (PT: $5 \times 10^{-08}$, $5 \times 10^{-07}$, $5 \times 10^{-06}$, $5 \times 10^{-05}$, .0005, .005, .05, .5 and 1). The PRS was standardised to have a mean of 0 and a standard deviation of 1; thus, a higher PRS represents higher genetic liability to each trait. We included SNPs that had a MAF of >1% and info score of >80% and excluded SNPs with an $R2$ of >0.1 if they were within 250 kb of each other. This was to account for linkage disequilibrium (LD) so that only the most strongly associated SNPs from each region were retained. Based upon previous research [78], we used the PRS with a p-value threshold of 1 in our analysis. | | | | | |
| Polygenic risk score for depression (see | NA | Child | Standardised to have a mean of | Birth | See genotyping information and genomic QC. |

| | | | | | |
|---|---|---|---|---|---|
| Howard et al. [12] for more details) | | | 0 and standard deviation of 1. | | |
| <u>Sleep health</u><br>Self-report sleep health items assessed at 'F@24' clinic at age 24 | | | | | |
| Sleep problems in past month | FKDQ4000 | Child | No (1), Yes (2) | 24 years, clinic questionnaire | 'In past month, YP had problems getting to sleep or back to sleep' |
| Sleep problems in past month | FKDQ4100 | Child | No (1), I have slept more than usual but this is not a problem (2), Yes (3) | 24 years, clinic questionnaire | 'In past month, sleeping more than usual has been a problem' |
| <u>Financial problems</u><br>Derived / summed variable using financial problems questions from the 'Life at 22' and 'Life at 23+' questionnaires | | | | | |
| Financial problems in early twenties | YPB6190 | Child | Yes, affected me a lot (1), Yes, moderately affected (2), Yes, mildly affected (3), Yes, but didn't affect me at all (4), No, did not happen (5) | 22+ years, questionnaire | QF20: 'Since age 21, whether had major financial problems and affect this had' |
| Financial problems in early twenties | YPC2360 | Child | No, did not happen (0), Yes, but did not affect respondent at all (1), Yes, mildly affected (2), Yes, moderately affected (3), Yes, affected respondent a lot (4) | 23+ years, questionnaire | QK22: 'Respondent had major financial problems since they were 22 years old, and degree to which it affected them' |

| | | | | | |
|---|---|---|---|---|---|
| <u>Problem cannabis use risk in early twenties</u><br>Derived / summed variables using items from the Cannabis Abuse Screening Test (CAST) from the 'It's All About You 20+' and 'Life at 22+' questionnaires, and 'F@24' clinic data<br>Items dichotomised into 'No, low, or moderate risk of problem cannabis use' (0) or 'High risk of problem use' (1) see Piontek et al. [79] for more information | | | | | |
| Cannabis Abuse Screening Test (CAST) score | CCCU3330 | Child | 0-6 | 20+ years, questionnaire | CAST score derived based on variables measuring cannabis use in past 12 months |
| Cannabis Abuse Screening Test (CAST) score | YPB4475 | Child | 0-6 | 22+ years, questionnaire | CAST score derived based on variables measuring cannabis use in past 12 months |
| Cannabis Abuse Screening Test (CAST) score | FKCA1025 | Child | 0-6 | 24+ years, clinic questionnaire | CAST score derived based on variables measuring cannabis use in past 12 months |
| <u>Illicit substance use in early twenties</u><br>Counts of amount of illicit substances used from the 'It's all about you 20+' and 'Life at 22+' questionnaires | | | | | |
| Count of types of drugs YP has ever used | CCU3470 | Child | 0-7 | 20+ years, questionnaire | Count of types illicit substances YP has ever used |
| Number of illicit substances YP ever used | YPB4477 | Child | 0-10 | 22+ years, questionnaire | Number of illicit substances YP has ever used |
| Alcohol Abuse Diagnoses in early twenties<br>Derived variables – any DSM-IV diagnoses of alcohol abuse and total DSM-IV alcohol abuse scores using items from the 'It's All About You 20+' and 'Life at 22+' questionnaires, and 'F@24' clinic data<br>Respondents to meet 3 of 7 alcohol abuse endorsements to meet DSM-IV alcohol abuse disorder criteria (<3=no diagnosis, >=3 diagnosis) | | | | | |
| Diagnosis of DSM-IV Alcohol Abuse | NA | Child | No diagnosis (0), Diagnosis (1) | 20+ years, questionnaire | Derived from alcohol use variables at age 20+ |
| Diagnosis of DSM-IV Alcohol Abuse | NA | Child | No diagnosis (0), Diagnosis (1) | 22+ years, questionnaire | Derived from alcohol use variables at age 22+ |
| Diagnosis of DSM-IV Alcohol Dependence | NA | Child | No diagnosis (0), Diagnosis (1) | 24+ years, clinic questionnaire | Derived from clinic alcohol use variable at age 24+ |

| | | | | | |
|---|---|---|---|---|---|
| Not in education, employment, or training (NEET) status in adolescence and early twenties<br>Derived variable used summing amount of NEET events experienced by respondent between the ages of 21 and 23 using 'Your Life Now' (21+), 'Life at 22+', and 'Life at 23+' questionnaires | | | | | |
| NEET status | YPA8000 | Child | Yes (1), No (2) | 21+ years, questionnaire | 'YP is currently in employment or doing any education or training' |
| NEET status (any events and total events) | NA | Child | Yes (1), No (0) | 22+ years, questionnaire | Derived from variables related to employment, education and training at age 22+ |
| NEET status (any events and total events) | NA | Child | Yes (1), No (0) | 23+ years, questionnaire | Derived from variables related to employment, education and training at age 23+ |
| Depression episodes from ages 23 to 28<br>Presence of depression episodes (as measured by Short Mood and Feelings Questionnaire (SMFQ)) from 'Life at 23+', 'Life at 25+' and 'Life at 28+' questionnaires | | | | | |
| Depression episodes between ages of 23 and 28 | NA | Child | Yes (1), No (0) | Amount of depression episodes derived from 23+, 25+, 28+ years questionnaires | Episodes defined as an SMFQ score >= 11, target variable derived from dichotomising whether respondent has experienced at least one episode of depression symptoms between the ages of 23 and 28 |

*Participant demographics – Models AF, HBL, CTAE*

| Variable | | Included in analysis (%) | Excluded from analysis (%) | $\chi 2$ | *P* |
|---|---|---|---|---|---|
| | **Participant sex (n=15,614)** | | | | |
| Males | | 34.93 | 53.31 | 663.17 | <0.001 |
| Females | | 65.07 | 41.87 | | |
| Not known | | 0 | 4.81 | | |
| | **Maternal education (n=12,470)** | | | | |
| A-level or higher | | 49.95 | 30.28 | 527.47 | <0.001 |
| O-level | | 33.73 | 34.94 | | |
| <O-level | | 16.31 | 34.78 | | |
| | **Maternal socioeconomic status (n=10,100)** | | | | |
| Professional / managerial / technical | | 46.91 | 33.73 | 149.71 | <0.001 |
| Skilled non-manual or lower | | 53.09 | 66.27 | | |
| | **Parity (n=13,101)** | | | | |
| First born | | 47.88 | 43.77 | 28.71 | <0.001 |
| Second born | | 34.91 | 34.94 | | |
| Third born or later | | 17.22 | 21.30 | | |
| | **Maternal age at pregnancy (n=14,269)** | | | | |
| <25 | | 13.25 | 26.51 | 307.47 | <0.001 |
| 25-29 | | 33.17 | 30.84 | | |
| 30-34 | | 35.93 | 24.55 | | |
| >35 | | 17.65 | 18.10 | | |

*Table 2: Demographic information for models AF, HBL and CTAE, demonstrating demographic differences between participants who had sufficient data to be included in the modelling versus those excluded.*

*Participant demographics – Model BF*

| Variable | Included in analysis, n (%) | Excluded from analysis, n (%) | $\chi 2$ | *P* |
|---|---|---|---|---|
| **Participant sex (n=15,614)** | | | | |
| Males | 38.23 | 49.21 | 170.74 | <0.001 |
| Females | 61.77 | 45.56 | | |
| Not known | 0 | 4.12 | | |
| **Maternal education (n=12,470)** | | | | |
| A-level or higher | 56.02 | 32.60 | 385.23 | <0.001 |
| O-level | 32.65 | 34.89 | | |
| <O-level | 11.33 | 32.51 | | |
| **Maternal socioeconomic status (n=10,100)** | | | | |
| Professional / managerial / technical | 53.68 | 34.99 | 166.55 | <0.001 |
| Skilled non-manual or lower | 46.32 | 65.01 | | |
| **Parity (n=13,101)** | | | | |
| First born | 50.73 | 44.03 | 37.75 | <0.001 |
| Second born | 34.57 | 34.97 | | |
| Third born | 14.70 | 21 | | |
| **Maternal age at pregnancy (n=14,269)** | | | | |
| <25 | 10.32 | 24.89 | 259.50 | <0.001 |
| 25-29 | 36.47 | 30.85 | | |
| 30-34 | 41.44 | 25.64 | | |
| >35 | 11.77 | 18.63 | | |

*Table 3: Demographic information for model BF, demonstrating demographic differences between participants who had sufficient data to be included in the modelling versus those excluded.*

*Table of performance evaluation metrics*

| Evaluation metric | Definition |
|---|---|
| Precision | Of the class predictions the model made, how many of the predictions were correct |
| Recall | Of the people who belonged to a particular class, how many of them did the model correctly identify |
| Macro F1-score | The harmonic mean between precision and recall |
| Area-under-the-curve (AUC) | The ability of the model to detect true positives and true negatives over false positives and false negatives |

*Table 4: Description of evaluation metrics for the random forest classifier.*
